# Interleukin-6 -174 G/C promoter gene polymorphism and polycystic ovary syndrome: A cross sectional investigation in adult women

**DOI:** 10.1101/2023.12.04.23299417

**Authors:** A Gupta, V Gupta, K Shah, V Gupta

## Abstract

Polycystic ovarian syndrome (PCOS), a metabolic disorder, manifests itself in a variety of ways. In this cross-sectional study, we evaluated the IL-6-174 G/C promoter gene polymorphism in adult PCOS women and its relationship to circulating levels and metabolic risk indicators. A total of 298 women between the ages between 15 and 35 years were chosen, 126 of whom had PCOS, and 172 of whom did not (control group). Both groups were further divided into subgroups of obese and lean women. The lipid profile, serum IL-6 level, and Homeostatic Model Assessment (HOMA) index were all examined. By using PCR-RFLP, the genotype of IL-6-174 G/C was identified. Women in the PCOS and non-PCOS groups showed significant variations in genotype frequencies and metabolic risk indicators. In PCOS compared to non-PCOS, there was a stronger correlation between the mutant ‘C’ allele of IL-6-174 G/C (p<0.0001; OR=1.91; 95% CI=1.38-2.66). Women with PCOS (61.2%) were significantly more associated with both the homozygous CC and heterozygous GC genotypes of IL-6-174 G/C in respect of WHR>0.85 than non-PCOS women (59.2%). In PCOS women, the distribution of the mutant genotypes CC and GC of the IL-6-174 G/C gene was likewise significantly different from GG, with higher WHR (p=0.0191), HOMA index (p=0.031), and serum IL-6 level (p=0.0094). These findings imply that the IL-6-174 G/C promoter mutant CC genotype was substantially related with increased circulating IL-6 levels, and that the presence of IR may be a risk indicator for the development of the metabolic syndrome (MetS) in PCOS women.

## 1. Introduction

A common heterogenous disorder known as polycystic ovary syndrome (PCOS) is characterized by hyperandrogenism, persistent anovulation, and infertility (*1*). According to World Health Organization (WHO) data, PCOS has an 8-13% prevalence worldwide and is one of the endocrine illnesses that affect women of reproductive age most frequently. Additionally, PCOS is a complicated metabolic disorder with an elevated lifetime risk of both diabetes and cardiovascular diseases (CVD) (*2*), although it is not a condition associated with insulin resistance (IR) or obesity. A high risk for impaired glucose tolerance and type 2 diabetes (T2D) has been shown in several investigations to be associated with PCOS (*3,4*). It is still unclear if this increased risk is a result of the anthropometrical and metabolic abnormalities typically found in PCOS or if it is connected to endocrine abnormalities such the hyperandrogenemia associated with PCOS.

IR is a common occurrence in women with PCOS, and it is often accompanied by compensatory hyperinsulinemia. Hyperinsulinemia appears to play a key pathogenic role in ovarian androgen overproduction because of the stimulatory effect of insulin on ovarian steroid production (*5*). The IR caused by hyperinsulinemia is intrinsic to PCOS and is not well understood. PCOS-specific IR affects both obese and lean women with PCOS. 50–60% of women with PCOS are obese, and they additionally possess a form of IR that is directly associated with obesity (*6*). Obesity, IR, and dyslipidemia are becoming more prominent risk indicators for metabolic syndrome (MetS) and CVD. In addition to obesity and IR, PCOS has been linked to elevated inflammatory markers associated with atherosclerosis disease, and genes coding for inflammatory cytokines are therefore considered candidates for a predisposition to risk of coronary heart disease and have been identified as target genes for PCOS (*7*).

Even fewer studies have concurrently taken into account several markers, such as interleukin-6 (IL-6), C-reactive protein (CRP), and tumor necrosis factor-α (TNF-α), when examining the prognostic value of inflammatory markers on CVD (*8, 9*). In comparison to CRP or TNF-α, IL-6 level has a more significant impact in the prediction of congestive heart failure (CHF) in older adults without a history of myocardial infarction (*8*). It is thought that IL-6 affects the process of fertilization and implantation because it possesses cytokine, endocrine, paracrine, and autocrine effects (*10*). Some of the PCOS-afflicted women had elevated levels of circulating IL-6 (*11*). Critical stages of follicular maturation are modified by an altered IL-6 level, which ultimately results in ovarian dysfunction (*12*). Thus, IL-6 may establish a connection between hyperandrogenemia in PCOS and anthropometric and metabolic abnormalities. The multifunctional cytokine IL-6 has a frequent G/C single nucleotide polymorphism at position 174 that has been shown to affect transcription rate (*13*) and is thought to be involved in the control of testosterone levels (*14*), insulin sensitivity, and energy expenditure (*15*). It has been suggested that people who carry the mutant IL-6 allele ‘C’ are more likely to experience lipid abnormalities, IR, and other PCOS-related disorders (*16*).

We aimed to investigate the association of IL-6-174 G/C gene polymorphism with IL-6 level and IR in PCOS women from North India. A second objective was to determine if correlations exist between IL-6-174 G/C promoter gene polymorphism and metabolic risk indicators associated with the MetS in women with PCOS.

## 2. Material and Methods

A total of 298 female participants from the Department of Obstetrics & Gynecology at Queen Mary Hospital, CSMMU, Lucknow, with ages ranging from 15 to 35 years were registered for the case-control research. Women were classified as obese (central obesity) if their WHR was greater than 0.85, and lean if it was less than 0.85. Out of them, 126 PCOS women (70 obese and 56 lean) were selected as the study group and the remaining 172 women (88 obese and 84 lean) who were non-PCOS (without PCOS) were chosen as the control group. PCOS was diagnosed using the revised criteria for PCOS following the 2003 Rotterdam consensus (*17*). The inclusion criteria for the women were that they had to be non-alcoholic, non-diabetic, and free of any form of metabolic, endocrine, cardiac, pulmonary, or inflammatory disease. The study excluded women who were pregnant, lactating, had any gynecological or obstetrical issues, were on medication, including hormone replacement treatment, or all of the above. The King George’s Medical University in Lucknow’s institutional ethics committee approved the study and and “all applicable institutional and governmental regulations concerning the ethical use of human volunteers were followed during this research”. Written informed consent was obtained from all participants.

In the first 10 days following the start of menstruation, all the women were examined for both normal menstruation and oligomenorrhea. A questionnaire was used to gather data on menstrual, nutritional, personal, familial, and medical histories. general examination; measurements of the waist to hip ratio (WHR) and body mass index (BMI). WHR (central obesity) was determined by measuring the hip circumference and abdomen circumference at the greater trochanter. With a 95% Confidence Interval (CI), 5% anticipated error, and 8% prevalence, sample size was determined by a power analysis.

### 2.1. Blood collection, biochemical analysis and DNA extraction

Each individual provided a total of 6.0 ml of venous blood, which was taken in the morning following a 12-hour fast. This blood sample was used to determine the serum biochemical and lipid profiles. 2.0 ml of that was taken and placed in EDTA vials. Blood was used to extract DNA using a commercial kit (Qiagen, USA). Plasma and serum were extracted from the remaining 4.0 ml of blood. The GOD-POD method was used to estimate the plasma Glucose (Randox Laboratories Ltd., Antrim, UK). Serum IL-6 was evaluated by ELISA (Quantikine IL-6, R&D system Oxford, UK), lipid profile by enzymatic method (Randox Laboratories Ltd., Antrim, UK), and serum insulin was assessed by immuno-radiometric assay method (Immunotech Radiova, Prague). By using the homeostatic model assessment index (HOMA Index), IR was estimated (*18*) using the equation:

HOMA Index = [fasting Insulin (μU/I) x fasting glucose (mmol/l)/ 22.5]

The HOMA Index 3.6 (*19*) was used to make a laboratory diagnosis of IR in people who did not meet the clinical and biological criteria for the condition.

### 2.2. Analysis of the IL-6-174 G/C gene

According to Fishman et al. (*13*), the genotyping procedure for the detection of the IL-6-174 G/C gene polymorphism was examined. All mutant and heterozygous samples were re-genotyped in order to increase the genotyping quality and validation, and only the results for reproducible and error-free samples were documented.

### 2.3. Statistical Analysis

Version 7 of the program GraphPad Prism was used to conduct statistical analysis. The mean and SD are used to express results. Odds ratios (OR) and their 95% confidence intervals were calculated using conditional logistic regression analysis to assess the relationship between the IL-6-174 G/C genotype and various metabolic risk indicators. Using gene counting and Fischer’s exact testing, the genotype and allele frequency of women with PCOS and those without were compared. By using ANOVA or an unpaired t test, the statistical significance of the difference between the means of metabolic risk indicators divided by the IL-6-174 G/C genotype was evaluated. Statistical significance was defined as a value of p<0.05.

## 3. Results

Based on 126 PCOS and 172 non-PCOS women between the ages of 15 and 35, or 25±10 years, the current study was conducted.

### 3.1. Metabolic risk indicators are compared between PCOS and non-PCOS subgroups of women

There was shown to be a significant difference between PCOS and non-PCOS women for the majority of the metabolic risk indicators. Those with PCOS had statistically higher Insulin (p=0.0033), HOMA Index (HOMA-IR) (p=0.0002), circulatory IL-6 level (p<0.0001), and total cholesterol (TC)/HDL ratio (p=0.0021) than those without PCOS or non-PCOS (Table 1). Without PCOS obese women (n=88, WHR>0.85) had high values for the HOMA Index (3.39±1.17 vs. 1.77±0.81, p<0.0001), TC/HDL (3.95±1.24 vs. 2.96±0.91, p<0.0001), Insulin (14.17±4.7 vs. 8.4±3.38, p<0.0001), serum triglyceride (TG) (133.45±30.04 vs. 118.95±30.56, p=0.0020), and circulatory IL-6 level (3.45±1.15 vs. 3.16±1.05, p=0.0834) than non-PCOS lean having WHR<0.85 (n=84). Similar results were seen for the HOMA Index (IR) (4.36±2.72 vs. 2.32±1.7, p<0.0001), TC/HDL (4.04±0.89 vs. 3.68±1.15, p=0.0498), Insulin (16.0±8.42 vs. 9.86±6.86, p<0.0001) and circulatory IL-6 level (11.04±2.71 vs. 5.96±1.55, p<0.0001) in PCOS obese women compared to PCOS lean women. Serum TG levels, however, showed no statistically significant difference (134.06±36.65 vs. 123.03±39.89, p=0.1091) (Table 1). With the exception of the HOMA Index (HOMA-IR) (4.36±2.72 vs. 3.39±1.17, p=0.0030) and circulatory IL-6 level (11.04±2.71 vs. 3.45±1.15, p<0.0001), there were no significant differences in serum TG and TC/HDL between PCOS obese and non-PCOS obese women (Figure 1).

**Table 1.** Metabolic risk indicators in women with PCOS, without PCOS, and in subgroups (lean and obese). (Unpaired student’s t-test)

| **Metabolic risk indicators** | **non-PCOS women (n=172)** | **PCOS women (n=126)** | **p- value** | **OR (95% CI)** |
| --- | --- | --- | --- | --- |
| Insulin | 10.93 ± 5.29 | 13.27 ± 8.32 | 0.0033* | 2.34 (0.79-3.89) |
| HOMA Index | 2.6 ± 1.3 | 3.5 ± 2.5 | 0.0002* | 0.85 (0.41-1.29) |
| TC/HDL | 3.47 ± 1.19 | 3.88 ± 1.03 | 0.0021* | 0.41 (0.15-0.67) |
| S. TG (mg/dl) | 123.3 ± 32.04 | 130.51 ± 39.55 | 0.4944 | 2.75 (5.16-10.66) |
| S. IL-6 (pg/ml) | 3.26 ± 1.14 | 8.91 ± 3.42 | <0.0001* | 5.65 (5.1- 6.2) |
| **Metabolic risk indicators** | **non-PCOS lean WHR <0.85 (n =84)** | **non-PCOS obese WHR >0.85 (n= 88)** | **p- value** | **OR (95% CI)** |
| Insulin | 8.4 ± 3.38 | 14.17 ± 4.7 | <0.0001* | 5.77 (4.53-7.0) |
| HOMA Index | 1.77 ± 0.81 | 3.39 ± 1.17 | <0.0001* | 1.6 (1.29-1.91) |
| TC/HDL | 2.96 ± 0.91 | 3.95 ± 1.24 | <0.0001* | 0.99 (0.66-1.32) |
| S. TG (mg/dl) | 118.95 ± 30.56 | 133.45 ± 30.04 | 0.0020* | 14.5 (5.38-23.62) |
| S. IL-6 (pg/ml) | 3.16 ± 1.05 | 3.45 ± 1.15 | 0.0834 | 0.29 (0.04-0.62) |
| **Metabolic risk indicators** | **PCOS lean WHR <0.85 (n =56)** | **PCOS obese WHR >0.85 (n= 70)** | **p- value** | **OR (95% CI)** |
| Insulin | 9.86 ± 6.86 | 16.0 ± 8.42 | <0.0001* | 6.14 (3.38-8.89) |
| HOMA Index | 2.32 ± 1.7 | 4.36 ± 2.72 | <0.0001* | 2.1 (1.28-2.92) |
| TC/HDL | 3.68 ± 1.15 | 4.04 ± 0.89 | 0.0498* | 0.36 (0.0003-0.72) |
| S. TG (mg/dl) | 123.03 ± 39.89 | 134.06 ± 36.65 | 0.1091 | 11.03 (2.49-24.56) |
| S. IL-6 (pg/ml) | 5.96 ± 1.55 | 11.04 ± 2.71 | <0.0001* | 5.08 (4.28-5.89) |

PCOS, polycystic ovary syndrome; HOMA, homeostatic model assessment; TC, total cholesterol; HDL, high density lipoprotein; TG, triglyceride; IL-6, interleukin-6; WHR, waist-hip-ratio; OR, odd ratio; CI, confidence interval
Data are presented as mean ±SD
Statistics were judged significant at p<0.05

**Figure 1.**
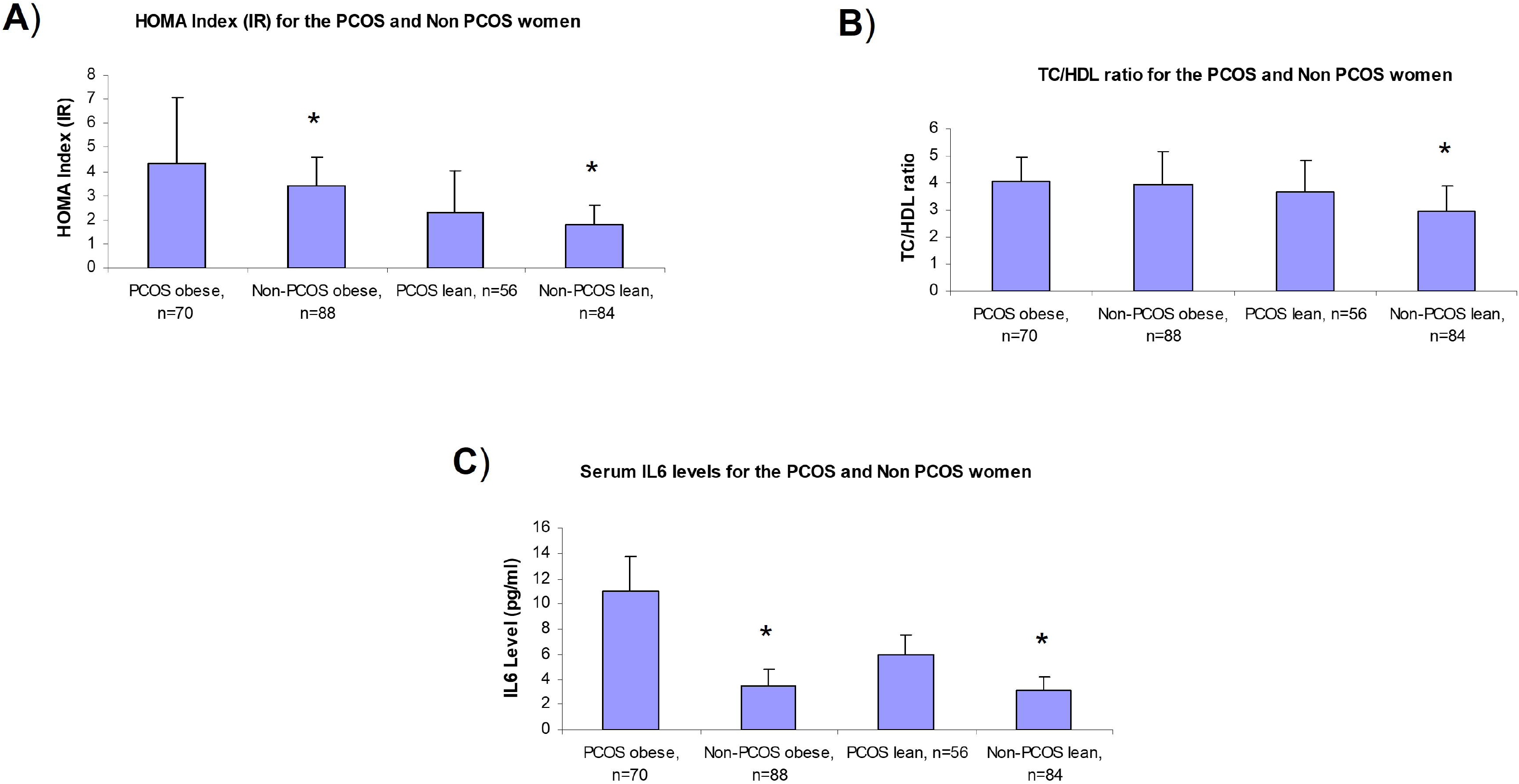
Metabolic risk indicators for women with PCOS, without PCOS, and for subgroups (Lean & Obese). A) HOMA Index, B) TC/HDL ratio, and C) Serum IL-6 level * p<0.05 for PCOS obese, non-PCOS obese, PCOS lean and non-PCOS lean subgroups

### 3.2. Distribution of the IL-6-174 G/C gene polymorphism in women with and without PCOS, including subgroups

We attempted to determine the distribution of the IL-6-174 G/C gene polymorphism in PCOS and non-PCOS women. When we compared these women, we discovered that non-PCOS women had a considerably greater frequency of the GG genotype (OR=2.35; CI=1.39-3.94, p=0.0012). The wild-type G allele was present in 42.1% and 58.1%, respectively, while the mutant C allele was present in 57.9% and 41.9%, respectively, in PCOS than non-PCOS women (Table 2). Additionally, it was discovered that PCOS women were substantially more likely to have the ‘C’ allele (OR=1.91, 95% CI= 1.38-2.66; p=0.0001) than non-PCOS women to have the ‘G’ allele.

**Table 2.** Distribution of the IL-6-174 G/C gene polymorphism among PCOS and non-PCOS women, and in subgroups (lean and obese)

| **IL-6-174**<br>**G/C gene** | **non-PCOS**<br>**women (n=172)**<br>**Alleles (n= 344)** | **PCOS women**<br>**(n=126)**<br>**Alleles (n= 252)** | **p- value** | **OR (95%CI)** |
| --- | --- | --- | --- | --- |
| **Genotypes** |  |  |  |  |
| GG | 69 (40.1 %) | 28 (22.2 %) | 0.0012* | 2.35 (1.39-3.94) |
| GC | 62 (36.1 %) | 50 (39.7 %) | 0.5464 | 0.86 (0.53-1.38) |
| CC | 41 (23.8 %) | 48 (38.1 %) | 0.0102* | 0.51 (0.31-0.84) |
| **Alleles** |  |  |  |  |
| G | 200 (58.1 %) | 106 (42.1 %) | 0.0001* | 1.91 (1.38-2.66) |
| C | 144 (41.9 %) | 146 (57.9 %) |  |  |
| **IL-6-174**<br>**G/C gene** | **PCOS lean**<br>**women (n=56)**<br>**Alleles (n=112)** | **PCOS obese women**<br>**(n=70)**<br>**Alleles (n=140)** | **p- value** | **OR (95%CI)** |
| **Genotypes** |  |  |  |  |
| GG | 18 (32.1 %) | 10 (14.3 %) | 0.0191* | 2.84 (1.19-6.81) |
| GC | 23 (41.1 %) | 27 (38.6 %) | 0.1408 | 1.75 (0.84-3.65) |
| CC | 15 (26.8 %) | 33 (47.1 %) | 0.0265* | 0.41 (0.19-0.87) |
| **Alleles** |  |  |  |  |
| G | 59 (52.7 %) | 47 (33.6 %) | 0.0031* | 2.20 (1.32-3.67) |
| C | 53 (47.3 %) | 93 (66.4 %) |  |  |
| **IL-6-174**<br>**G/C gene** | **non-PCOS lean**<br>**women (n=84)**<br>**Alleles (n=168)** | **non-PCOS obese**<br>**women (n=88)**<br>**Alleles (n=176)** | **p- value** | **OR (95%CI)** |
| **Genotypes** |  |  |  |  |
| GG | 41 (48.8 %) | 28 (31.8 %) | 0.0293* | 2.04 (1.09-3.79) |
| GC | 27 (32.1 %) | 35 (39.8 %) | 0.3418 | 0.72 (0.38-1.34) |
| CC | 16 (19.1 %) | 25 (28.4 %) | 0.1577 | 0.59 (0.29-1.21) |
| **Alleles** |  |  |  |  |
| G | 109 (64.9 %) | 91 (51.7 %) | 0.0161* | 1.73 (1.12-2.66) |
| C | 59 (35.1 %) | 85 (48.3 %) |  |  |

PCOS, polycystic ovary syndrome; IL-6, interleukin-6; WHR, waist-hip-ratio; OR, odd ratio; CI, confidence interval
Statistics were judged significant at $p < 0.05$

It was also discovered that the prevalence of the GG genotype was substantially greater in PCOS lean women compared to PCOS obese women (OR= 2.84; CI=1.19-6.81, p=0.0191). The wild-type G allele was present in 33.6% of obese and 52.7% of lean PCOS women, but the mutant C allele was present in 66.4% and 47.3%, respectively (Table 2). Additionally, it was discovered that the ‘C’ allele was more strongly related with PCOS obese women than PCOS lean women (OR=2.20, 95% CI= 1.32-3.67; p=0.0031) than the ‘G’ allele.

non-PCOS lean women were shown to have a considerably greater frequency of the GG genotype (OR=2.04; CI=1.09-3.79, p=0.0293) than non-PCOS obese women. The wild-type G allele was present in 51.7% and 64.9% of non-PCOS obese and non-PCOS lean women, respectively, while the mutant C allele was present in 48.3% and 35.1% of both groups, respectively. Additionally, it was discovered that the ‘C’ allele was more strongly related with non-PCOS obese women than the ‘G’ allele was with lean women (OR=1.73, 95% CI= 1.12-2.66; p=0.0161) (Table 2).

### 3.3. IL-6-174 G/C gene polymorphism prevalence as determined by metabolic risk indicators

We attempted to connect the existence of metabolic risk indicators in PCOS and non-PCOS women with the IL-6-174 G/C promoter gene polymorphism (Table 3). When compared to the wild type GG genotype of the IL-6 gene, it has been found that mutant, homozygous CC as well as heterozygous GC are more strongly related with high WHR >0.85 (p=0.0191), HOMA Index >3.6 (p=0.031), and circulatory IL-6 level >2.5 pg/ml (p=0.0094). However, no statistically significant correlations were discovered for serum TG >150 mg/dl (p=0.6126) and TC/HDL >3.8 (p=0.3961). However, the relationship between the IL-6-174 G/C polymorphism and anthropometric variables and metabolic risk indicators in non-PCOS women revealed a significant association between high WHR>0.85 (p=0.0001), HOMA Index >3.6 (p=0.0001), and TC/HDL>3.8 (p=0.0001) with homozygous (CC) and heterozygous (GC) mutation as compared to wild type GG genotype of the IL-6 gene. However, no statistically significant correlation was detected between any type of mutation and serum TG levels >150 mg/dl (p=0.2987) and circulatory IL-6 level >2.5 pg/ml (p=0.8702) (Table 3).

**Table 3.** IL-6-174 G/C gene polymorphism prevalence among PCOS and non-PCOS women (GG vs. GC + CC genotype) in relation to metabolic risk indicators.

| Metabolic risk indicators | GG (n=28) | CC + GC (n=98) | p- value | OR (95% CI) |
| --- | --- | --- | --- | --- |
| **PCOS women (n=126)** |  |  |  |  |
| WHR (>0.85) | 10 (35.7 %) | 60 (61.2 %) | 0.0191* | 0.44 (0.22-0.89) |
| HOMA Index (>3.6) | 07 (25.0 %) | 48 (48.9 %) | 0.031* | 0.43 (0.19-0.94) |
| TC/HDL (>3.8) | 15 (53.6 %) | 43 (43.9 %) | 0.3961 | 0.68 (0.29-1.58) |
| S. TG (>150 mg/dl) | 05 (17.9 %) | 24 (24.5 %) | 0.6126 | 1.49 (0.51-4.36) |
| S. IL-6 (>2.5 pg/ml) | 16 (57.1 %) | 81 (82.7 %) | 0.0094* | 0.39 (0.21-0.74) |
| Metabolic risk indicators | GG (n=69) | CC + GC (n=103) | p- value | OR (95% CI) |
| **non-PCOS women (n=172)** |  |  |  |  |
| WHR (>0.85) | 17 (24.6 %) | 61 (59.2 %) | <0.0001* | 4.44 ( 2.26-8.72) |
| HOMA Index (>3.6) | 06 (8.7 %) | 39 (37.9 %) | <0.0001* | 6.39 ( 2.53-16.18) |
| TC/HDL (>3.8) | 12 (17.4 %) | 60 (58.3 %) | <0.0001* | 6.63 (3.17-13.81) |
| S. TG (>150 mg/dl) | 09 (13.1 %) | 21 (20.4 %) | 0.2987 | 1.71 (0.73-3.99) |
| S. IL-6 (>2.5 pg/ml) | 16 (23.2 %) | 25 (24.3 %) | 0.8702 | 1.06 (0.52-2.18) |

PCOS, polycystic ovary syndrome; WHR, waist-hip-ratio; HOMA, homeostatic model assessment; TC, total cholesterol; HDL, high density lipoprotein; TG, triglyceride; IL-6, interleukin-6; OR, odd ratio; CI, confidence interval
Statistics were judged significant at $p < 0.05$

## 4. Discussion

The most prevalent endocrine condition in females, PCOS has a mysterious pathophysiologic and molecular background. A recent meta-analysis of 11 studies showed that Indian women have higher prevalence of PCOS which is close to 10% according to the Rotterdam’s criteria *(20)*. One-third of PCOS-afflicted women exhibit central obesity as well as several CVD risk indicators, including glucose intolerance. IL-6, a cytokine mostly produced by adipose tissue, has just lately been linked to the emergence of cardiovascular events (*21,22*). Understanding of the genetic associates of these metabolic abnormalities can shed light on underlined pathophysiological mechanisms of the disease in specific population.

In the current study, we investigated the association between circulating IL-6 level, IL-6-174 G/C promoter gene polymorphism, and metabolic risk indicators in PCOS women. Our research showed that women with PCOS had higher levels of circulatory IL-6 than women without the condition, and that this rise in serum IL-6 was more closely linked to obesity in PCOS women. Similarly, obese women with PCOS and those without it had higher IR than lean women. Instead of PCOS itself, central fat excess is linked to an increase in serum IL-6 level and IR in PCOS women. Our findings are in line with those of past research (*23-25*) that found higher blood levels of inflammatory markers such TNF-α, IL-6, and hs-CRP in PCOS women. Additionally, in adult PCOS women, variations in IL-6 serum concentrations coincided with variations in both total and abdominal fat mass. These were linked to both obesity and IR, which are both frequent symptoms of PCOS (*26*).

In healthy people, adipose tissue contributes up to 30% of the total circulatory IL-6 concentration. WHR has been proven to correlate with T2D and provides a decent indication of visceral obesity (*27*). Our findings showed that, compared to PCOS women with wild-type GG genotype, those with mutant, homozygous CC and heterozygous GC genotype were more likely to have high WHR >0.85 (p=0.0191), HOMA index (p=0.031), and circulating IL-6 level >2.5 pg/ml (p=0.0094). Similarly, higher values for WHR >0.85, HOMA Index >3.6, and serum TC/HDL-C >3.8 were substantially related with non-PCOS obese women with the mutant homozygous CC and heterozygous GC genotypes. Therefore, the presence of the mutant C allele of the IL-6-174 G/C promoter gene as opposed to its wild type G allele was more strongly connected with the existence of metabolic risk indicators like central obesity, IR, dyslipidemia, and high serum levels of IL-6. Less evidence, though, supports the idea that IL-6 affects insulin function. In response to IL-6 treatment, one recent study found that adipocyte IRS-1 (insulin receptor substrate-1), GLUT4 expression, and tyrosine phosphorylation decreased (*28*) while other research points to the possibility that IL-6 may stimulate fat oxidation in adipocytes, increasing the levels of circulating NEFAs (*29*).

Comparing PCOS women to non-PCOS women who were age and BMI matched, our study found no evidence of a significant difference in serum TG levels. Others have demonstrated that PCOS patients exhibited dyslipidemia that was linked to insulin-resistance states and were more insulin resistant (*30*). Even after controlling for age and total body fat, they still reported higher serum levels of a number of inflammatory markers. A more centralized fat buildup was used to explain this PCOS impact (*31*). Due to the cross-sectional nature of the study, a directionality of cause and effect between obesity, IR, and circulatory IL-6 level cannot be conclusively demonstrated. However, it is clear that there are connections between metabolic risk indicators, IR, and obesity that must be looked at separately.

Compared to lean individuals, PCOS obese individuals demonstrated greater levels of circulatory IL-6 and IR and were more substantially related with the presence of metabolic risk indicators. In both PCOS and non-PCOS women, it was discovered that the mutant, homozygous CC genotype at the 174 G/C promoter region of the IL-6 gene was linked to higher levels of circulating IL-6, WHR, and IR. Women without PCOS who had high TC/HDL values also had higher levels of IL-6. Therefore, visceral obesity and the presence of mutant CC genotype in PCOS were linked more strongly than they were independently to the increase in metabolic risk indicators, including IR, serum IL-6, and high values of TC/HDL, responsible for the development of the MetS. Therefore, lifestyle changes and weight loss among PCOS women may significantly reverse the metabolic risk that goes along with it. This is important observation from a public health aspect of disease management as weight loss can be achieved majority of the times through non-pharmacological, cost-effective interventions. This can help patients in escaping the vicious cycle of weight gain, metabolic syndrome and then even more severe forms of metabolic diseases.

## Data Availability

All data produced in the present work are contained in the manuscript.

## 6. Author contribution statements

AG & VG are responsible for conception and design of the study. AG & VdG performed the experiments. AG & VG analyzed the data. AG, VdG & VG interpreted the results of the experiments. AG prepared the figure. AG, VdG, KS & VG drafted the manuscript. AG, VdG, KS and VG edited the content of the manuscript. All authors read and approved the final manuscript.

## 9. Acknowledgement

We appreciate the Queen Mary Hospital’s faculty and staff from King George’s Medical University for giving PCOS patients and the Department of Medical Genetics, SGPGI Lucknow for genetic analysis.

## FIGURE

**Figure 1**. Metabolic risk indicators for women with PCOS, non-PCOS, and for subgroups (Lean & Obese): A) HOMA Index (IR) level, B) TC/HDL ratio, and C) Serum IL-6 level (pg/ml)

PCOS, polycystic ovary syndrome; HOMA, homeostatic model assessment; TC, total cholesterol; HDL, high density lipoprotein; IL-6, interleukin-6

Statistics were judged significant at *p<0.05; PCOS obese vs. non-PCOS obese, PCOS lean vs. non-PCOS lean

